# Clinical and Paraclinical Characteristics of COVID-19 patients: A systematic review and meta-analysis

**DOI:** 10.1101/2020.03.26.20044057

**Authors:** Keyvan Heydari, Sahar Rismantab, Amir Shamshirian, Parisa Lotfi, Nima Shadmehri, Pouya Houshmand, Mohammad Zahedi, Danial Shamshirian, Sahar Bathaeian, Reza Alizadeh-Navaei

## Abstract

**Introduction:** Recently, a new strain of coronaviruses, which originated from Wuhan City, Hubei Province, China has been identified. According to the high prevalence of new coronavirus, further investigation on the clinical and paraclinical features of this disease seems essential. Hence, we carried out this systematic review and meta-analysis to figure out the unknown features.

**Methods:** This study was performed using databases of Web of Science, Scopus and PubMed. We considered English cross-sectional and case-series papers which reported clinical, radiological, and laboratory characteristics of patients with COVID-19. We used STATA v.11 and random effect model for data analysis.

**Results:** In the present meta-analysis, 32 papers including 49504 COVID-19 patients were studied. The most common clinical symptoms were fever (84%), cough (65%) and fatigue (42%), respectively. The most common radiological and paraclinical features were bilateral pneumonia (61%), ground-glass opacity (50%), thrombocytopenia (36%) and lymphocytopenia (34%). The study also showed that the frequency of comorbidities and early symptoms was higher in critically severe patients. Moreover, we found the overall mortality rate of three percent.

**Conclusion:** According to that there are many cases without Computed Tomography Scan findings or clear clinical symptoms, it is recommended to use other confirming methods such RNA sequencing in order to identification of suspicious undiagnosed patients. Moreover, while there is no access to clinical and paraclinical facilities in in public places such as airports and border crossings, it is recommended to consider factors such as fever, cough, sputum and fatigue.

## Introduction

Coronavirus is a type of enveloped RNA virus with size of 60 to 140 nanometers in diameter. They have spike-like projections with a crown-like appearance under the electron microscope (1). This family of viruses, which can be divided into four type of alpha, beta, delta, and gamma, can infect multiple types of different species. For instance, they have been isolated from humans (alpha and beta), bats, Canidae, and Felidae (2, 3). Although the coronavirus family is mainly responsible for human mild respiratory diseases such as common cold, in some cases, they can cause serious illnesses (3). Middle east respiratory syndrome (MERS) and severe acute respiratory syndrome (SARS) are among epidemic diseases caused by coronaviruses in recent decades (4).

Recently, a novel strain of coronaviruses, which originated from Wuhan City, Hubei Province, China, has been identified (1). This strain (SARS-CoV-2) has been discovered in 2019 and characterized as a pandemic disease in March 2020 (5). According to World Health Organization (WHO), up to 17 March 2020, over 188,000 confirmed cases of COVID-19 had been reported throughout the world with the mortality rate of 3.4 % (6).

In a study, Sun *et al*. assessed the incidence of fever, cough, myalgia, fatigue and ARDS in patients with CoVID-19 (7). Given the increasing prevalence of the disease, further investigation of the clinical and paraclinical features of this disease seems necessary. In this regard, we conducted this systematic and meta-analysis review. In this study, we investigated the risk of incidence of different type of clinical finding and comorbidities in severe and non-severe patients.

## Material and Methods

### Source information

This systematic review and meta-analysis has been carried out on cross-sectional and case-series studies. We searched databases of Web of Science, Scopus, and PubMed without any time limitation for publications up to March 13, 2020. As manual search, the list of imported references, list of related reviews, and the results of Google Scholar have been investigated.

### Search strategy

All processes of searching, screening and reporting were done according to Preferred Reporting Items for Systematic Reviews and Meta-Analyses (PRISMA) guideline. We used following keywords in our systematic search in order to find English papers: “2019 novel coronavirus infection”, COVID19, “coronavirus disease 2019”, “coronavirus disease-19”, “2019-nCoV disease”, “2019 novel coronavirus disease”, and “2019-nCoV infection.”; all in title/abstract field.

### Eligibility Criteria

We considered following criteria for study selection:

- The study should have an observatory approach investigated COVID-19 patients.
- Studies on a particular group of people (e.g. pregnant women) has been excluded.
- Studies included with desired variables such as symptoms, comorbidity, laboratory findings, etc.
- Studies should be in English. English abstracts of other language studies were investigated for eligible data.
- All the case report and animal studies were excluded.

### Study Selection

Duplicated papers were deleted using EndNote software. Two researchers screened the remaining papers according to the criteria, separately. In case of disagreement between the two investigators, a third person was making the final decision.

### Quality assessment

For the quality assessment, a modified version of Newcastle-Ottawa Quality Assessment (NOS) have been used. The investigated papers categorized into two categories. The studies with a score of 1 or 2 as poor quality, and studies with a score of 3 to 5 as high quality.

### Data Extraction

Data such as authors information, publication year, sample size, average/median age, death rate, hospitalization status, early symptoms, laboratory findings, radiological findings and underlying diseases of patients have been extracted and recorded.

### Statistical Analysis

Statistical analysis was performed using STATA v.11 software. The heterogeneity of studies has been investigated using I-square (*I*^*2*^) test. Based on the results, for *I*^*2*^ more than 50%, we used a random-effects model to pool the results. Moreover, to study the heterogeneity of the added study, subgroup analysis in severe and non-severe patients have been done.

## Results

### Study Selection Process

The search in databases yields 351 results. After excluding duplicated papers, 364 results were admitted to the screening step. Then, 65 papers have been selected for full-text analysis. Finally, 19 studies were entered into the meta-analysis. Additionally, 13 studies obtained from manual search were included in the meta-analysis. The PRISMA flow diagram for the study selection process presented in Figure 1.

**Figure 1.**
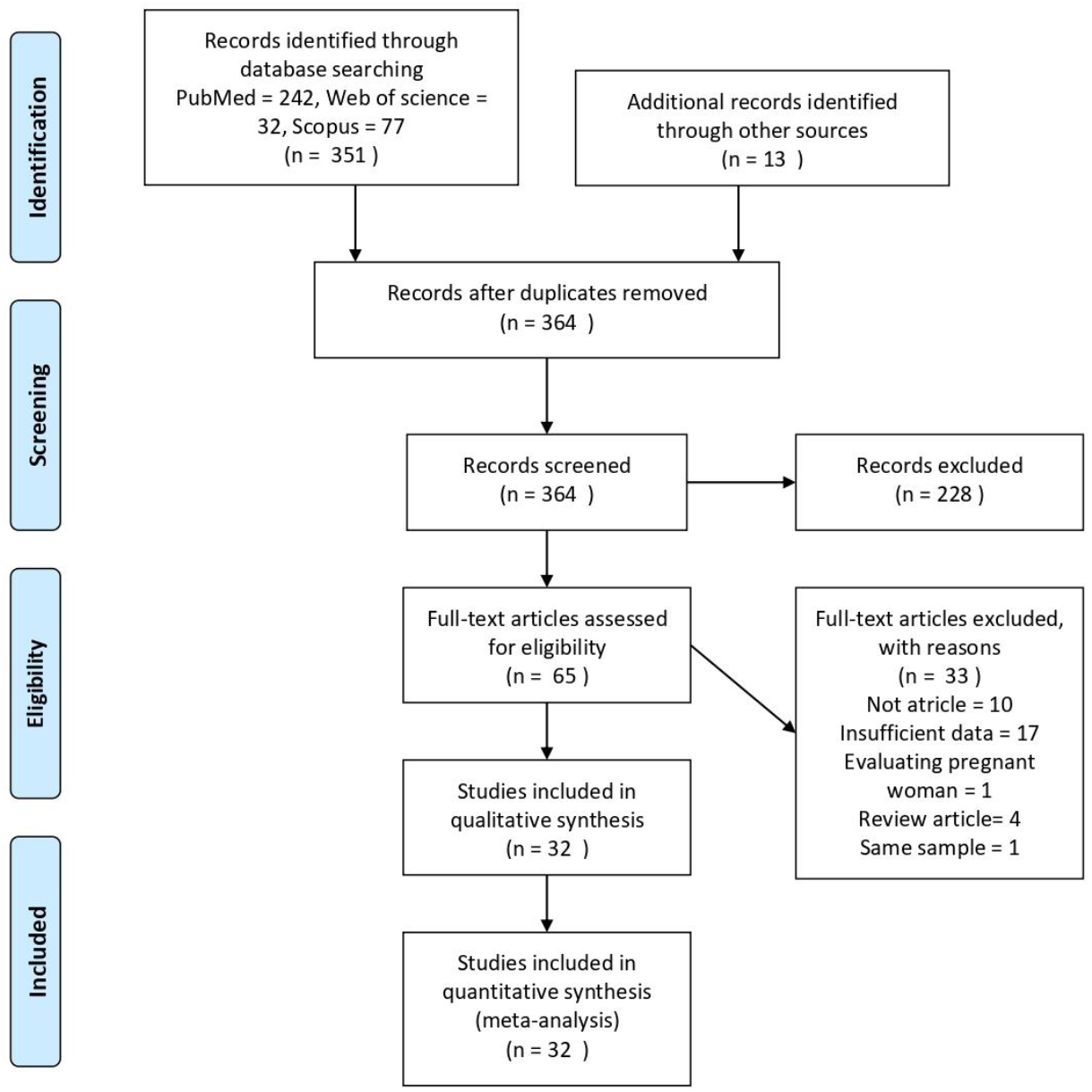
PRISMA flowchart for study selection process.

### Study Characteristics

Out of selected papers, a total of 49504 patients infected with CoVID-19 with age ranged between 40 to 58 years old were included in our investigation. All of the papers were conducted in China and only one study was a collaboration between China and South Korea. Characteristics of studies entered into meta-analysis are presented in Table 1.

**Table 1.** Characteristics of studies entered into the meta-analysis

| Author | Country | Type<br>of<br>study | No. of<br>Patients | Mean<br>Age<br>(SD) | Clinical<br>findings | WBC count<br>(10<sup>9</sup>/L) | Neutrophil<br>count<br>(10<sup>9</sup>/L) | Lymphocyte<br>count (10<sup>9</sup>/L) | Platelet count<br>(10<sup>9</sup>/L) | Comorbidities | Q/A<br>score |
| --- | --- | --- | --- | --- | --- | --- | --- | --- | --- | --- | --- |
| Ai (11) | China | R | 1014<br>(467/547) | 51 (15) | - | - | - | - | - | - | 3 |
| Jiehao (12) | China | CS | 10 (4/6) | - | Fever, Cough,<br>Sneezing, Nasal<br>congestion,<br>Rhinorrhea, Sore throat | - | - | - | - | - | 3 |
| Chen (A)<br>(13) | China | R | 99 (67/32) | 55.5<br>(13.1) | Fever, Myalgia, Cough,<br>Rhinorrhea, Sore<br>throat, Shortness of<br>breath, Confusion,<br>Headache, Chest pain,<br>Diarrhea, N/V | - | - | - | - | CVD, DD, ESD,<br>MD, CNSD,<br>RSD, | 4 |
| Chen (B)<br>(14) | China | R | 29 | - | Fever | - | - | - | - | - | 3 |
| Hu (15) | China | CS | 24 | - | - | - | - | - | - | - | 3 |
| Huang (16) | China | R | 41 (30/11) | 49 | Fever, Myalgia, Cough,<br>Sputum production,<br>Dyspnea, Headache,<br>Diarrhea | 6.2 (4.1-10.5) | 5 (33-8.9) | 0.8 (0.6-1.1) | - | CVD, DM,<br>HTN, DD, MD,<br>RSD, | 5 |
| Ki (17) | Korea/Chi<br>na | CS | 28 (15/13) | - | Fever, Myalgia,<br>Sneezing, Sputum<br>production, Chills,<br>Sore throat, Headache | - | - | - | - | - | 4 |
| Liu (A)<br>(18) | China | CS | 12 (8/4) | - | Fever, Myalgia, Cough,<br>Chills, Diarrhea, N/V | - | - | - | - | CVD, DM,<br>HTN, RSD,<br>CKD | 4 |
| Liu (B)<br>(19) | China | CS | 61 (31/30) | 40* | Fever, Cough, Sputum<br>production, Chills,<br>Sore throat, Shortness<br>of breath, Dyspnea,<br>Headache, Fatigue, | 4.3 (3.5-5.1) | 2.5 (2.1-3.5) | - | 164 (135.0-219.5) | CVD, DM,<br>HTN, RSD, | 4 |
|  |  |  |  |  | Chest pain, Diarrhea, |  |  |  |  |  |  |
|  |  |  |  |  | Anorexia, N/V |  |  |  |  |  |  |
| Qin (20) | China | CS | 452 (235/217) | 58* | Abdominal pain, | 5.3(3.9-7.5) | 3.9 (2.6-5.8) | 0.9 (0.6-1.2) | - | CVD, DM, CD, | 4 |
|  |  |  |  |  | Pharyngalgia, |  |  |  |  | HTN, CLD, |  |
|  |  |  |  |  | Hemoptysis, |  |  |  |  | MD, RSD, CKD |  |
|  |  |  |  |  | Expectoration, Fever, |  |  |  |  |  |  |
|  |  |  |  |  | Myalgia, Sneezing, |  |  |  |  |  |  |
|  |  |  |  |  | Dizziness, Rhinorrhea, |  |  |  |  |  |  |
|  |  |  |  |  | Shortness of breath, |  |  |  |  |  |  |
|  |  |  |  |  | Confusion, Headache, |  |  |  |  |  |  |
|  |  |  |  |  | Fatigue, Diarrhea, |  |  |  |  |  |  |
|  |  |  |  |  | Anorexia, N/V |  |  |  |  |  |  |
| Wu (A) | China | R | 80 (39/41) | 46.1 | Fever, Myalgia, Cough, | 4.1 (3.2-5.7) | 4.3 (2.3-5.9) | 0.6 (0.4-1.0) | 155 (116-188) | CVD, DD, | 5 |
| (21) |  |  |  | (15.42) | Rhinorrhea, Sore |  |  |  |  | CLD, ESD, |  |
|  |  |  |  |  | throat, Shortness of |  |  |  |  | MD, |  |
|  |  |  |  |  | breath, Headache, |  |  |  |  | CNSD,RSD, |  |
|  |  |  |  |  | Diarrhea, N/V |  |  |  |  | CKD |  |
| Wu (B) | China | R | 201 (128/73) | 51* | Fever, Cough, | 5.94 (3.80- | 4.47 (2.32- | 0.91 (0.60- |  | CVD, DM, | 3 |
| (22) |  |  |  |  | Dyspnea, Fatigue | 9.08) | 7.70) | 1.29) |  | HTN, CLD, |  |
|  |  |  |  |  |  |  |  |  |  | MD, RSD, ESD |  |
| Zhu (23) | China | R | 32 (15/16) | 46* | Expectoration, Fever,<br>Cough, Headache,<br>Fatigue, Diarrhea | - | - | - | 157.2 (83 - 284) | CVD, DM,<br>HTN, CLD,<br>MD, ESD, CD, | 4 |
| Xie (24) | China |  | 9 (4/5) | - | Fever, Cough, Fatigue,<br>Diarrhea | - | - | - | - | - | 4 |
| Xu (25) | China | CS | 62 (36/27) |  | Hemoptysis,<br>Expectoration,<br>Myalgia, Cough,<br>Headache, Chest pain,<br>Diarrhea | 4.7 (3.5-5.8) | 2.0 (2.0-3.7) | 1.0 (0.8-1.5) | 176 (135.8-215.5) | CVD, DM,<br>HTN, CLD,<br>RSD, | 5 |
| Yu (26) | China |  | 887 | - | - | - | - | - | - | - | 3 |
| Guan (27) | China | R | 1099<br>(640/459) | 47* | Hemoptysis, Fever,<br>Myalgia, Cough,<br>Sputum production,<br>Chills, Nasal<br>congestion, Sore throat,<br>Shortness of breath,<br>Headache, Fatigue,<br>Diarrhea, N/V | 4.7 (3.5-6) | - | - | 168 (132-207) | CVD, DM, CD,<br>HTN, MD,<br>RSD, ID, CKD, | 5 |
| Bernheim<br>(28) | China | CS | 121 (61/60) | 45<br>(15.6) | Fever, Cough, Sputum<br>production | - | - | - | - | - | 3 |
| Chung (29) | China | CS | 21(13/8) | 51<br>(14.5) | Fever, Cough, Myalgia,<br>Headache, Fatigue,<br>N/V | - | - | 1.0 (0.7-1.3) | - | - | 5 |
| Chang (30) | China | CS | 13 (10/3) | 34 | Myalgia, Cough, Nasal<br>congestion,<br>Rhinorrhea, Headache,<br>Diarrhea | 5.83 | 3.67 |  |  |  | 4 |
| Cheng (31) | China | CS | 11 (8/3) | 50..36<br>(15.5) | Fever, Cough, Myalgia,<br>Sputum production,<br>Sore throat, Shortness<br>of breath, Diarrhea | 4.38 (3.2–<br>5.01) | 3.13 (2.02–<br>3.67) | 1.10 (1.03–<br>1.33) | - |  | 4 |
| Pan (A)<br>(32) | China | R | 21 (6/15) | 40* | Expectoration, Fever,<br>Myalgia, Cough,<br>Chills, Fatigue, Chest<br>pain, Anorexia | 4.9 (3.1-6.9) | 3.1 (2.1(4.6) | 1.4 (0.7-2.5) | - | - | 4 |
| Pan (B)<br>(33) | China | CS | 63 (33/30) | 44.9<br>(15.2) | - | - | - | - | - | - | 4 |
| Yang (34) | China | R | 52 (35/17) | 59.7<br>(13.3) | Fever, Myalgia, Cough,<br>Rhinorrhea, Headache,<br>Chest pain, N/V | - | - | - | - | DM, CD, MD,<br>CNSD, RSD,<br>CKD | 3 |
| China<br>CDC<br>(35) | China | CS | 44672(22981/<br>21691) | - | - | - | - | - | - | - | 5 |
| Zhang (36) | China | CS | 9 (5/4) | 36* | Pharyngalgia, Myalgia,<br>Fever, Cough, , Nasal<br>congestion,<br>Rhinorrhea, Fatigue | - | - | - | - | - | 4 |
| Han (37) | China | CS | 108 (38/70) | 45* | Pharyngalgia, Fever,<br>Myalgia, Cough,<br>Headache, Chest pain,<br>Fatigue, Diarrhea |  |  |  |  |  | 3 |
| Feng (38) | China | CS | 15 (5/10) | - | - | - | - | - | - | - | 3 |
| Wang (A)<br>(39) | China | CS | 138 (75/63) | 56* | Abdominal pain,<br>Pharyngalgia,<br>Expectoration, Fever,<br>Myalgia, Cough,<br>Dizziness, Dyspnea, | 4.5 (3.3-6.2) | 3.0 (2.0-4.9) | 0.8 (0.6-1.1) | 163 (123-191) | CVD, DM, CD,<br>HTN, CLD,MD,<br>RSD, ID, CKD, | 5 |
|  |  |  |  |  | Headache, Fatigue,<br>Diarrhea, Anorexia,<br>N/V |  |  |  |  |  |  |
| Wang (B)<br>(40) | China | CS | 34 (14/20) | - | Pharyngalgia, Fever,<br>Cough | 3.82(2.98-5.57) | 2.35(1.62-367) | 1.15 (0.82 –<br>1.46) | 171 (142 – 211) | - | 4 |
| Wang (C)<br>(41) | China | R | 69 (32/37) | - | Pharyngalgia, Myalgia,<br>Cough, Dizziness,<br>Sputum production,<br>Dyspnea, Headache,<br>Fatigue, Diarrhea,<br>Anorexia, N/V, , Chest<br>pain | 3.82(2.98-5.57) | 2.35(1.62-367) | 1.15(0.82-1.46) | 171 (142 -211) | CVD, DM,<br>HTN, CLD,<br>MD, RSD | 4 |
| Li (42) | China | CS | 17 (9/8) | 45.1<br>(12.8) | Fever, Myalgia, Cough,<br>Sputum production,<br>Dizziness, Rhinorrhea,<br>Fatigue, Diarrhea | - | - | - | - | HTN, CLD,<br>RSD, | 4 |

\*: Median
R; retrospective, CS: Case series, N/V: Nausea/vomiting, CVD: cardiovascular disease, DM: Diabetes, CD: cerebrovascular disease, HTN: hypertension, DD: digestive disease, CLD: chronic liver disease, ESD: endocrine system disease, MD: malignant disease, CNSD: Central neurons system disorder, RSD: respiratory system disease, ID: immunodeficiency, CKD: chronic kidney disease,

### Quality Assessment

According to quality assessment using NOS tool, all of the studies categorized as high quality. The quality assessment graph is presented in Fig. 2 and Supplementary Fig. 1.

**Figure 2.**
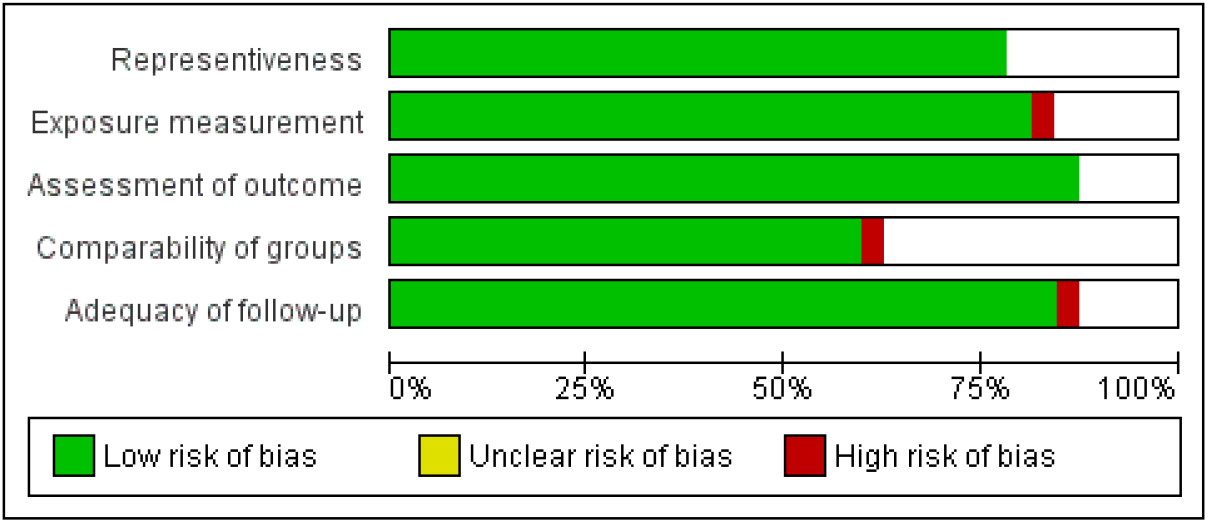
Risk of bias summary (A)

### Characteristics of Patients

Regarding hospitalization, our findings showed that 69% of the patients need to be hospitalized. About 26% of patients were discharged after receiving outpatient treatment and only 3% of patients have been expired (Fig. 3).

**Figure 3.**
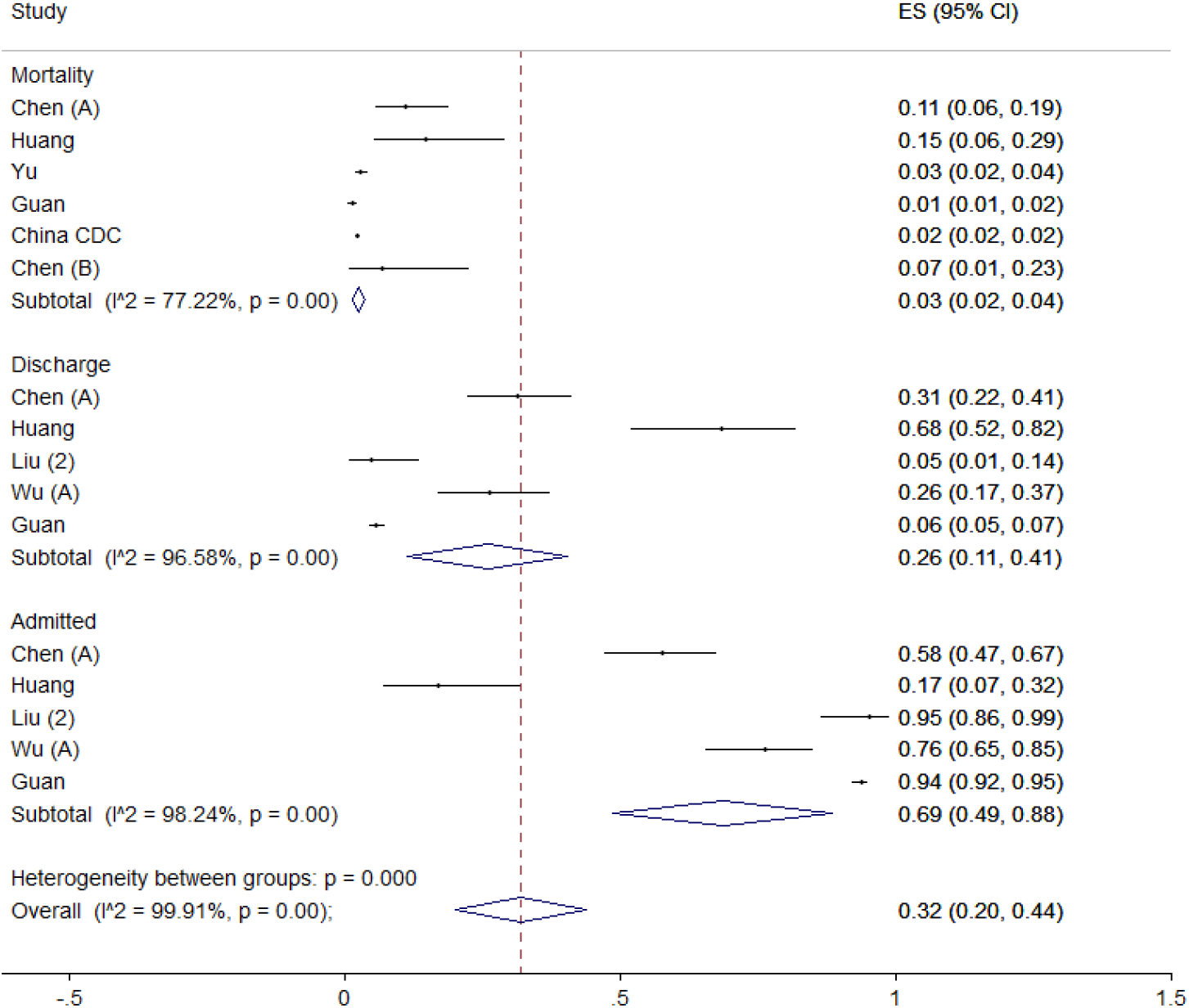
Meta-analysis of incidence of clinical outcome of patients.

### Clinical Findings

Patients with COVID-19 referred to the hospital with variety symptoms. Fever, cough, and sour throat are among the most common symptoms of the patients, which were reported respectively in 22, 20, and 7 studies. Meta-analysis findings are as follows respectively: 84% (95% CI, 79-88), 65% (95% CI, 59-71), and 14% (95% CI 8-19) (Fig. 4 and 5).

**Figure 4.**
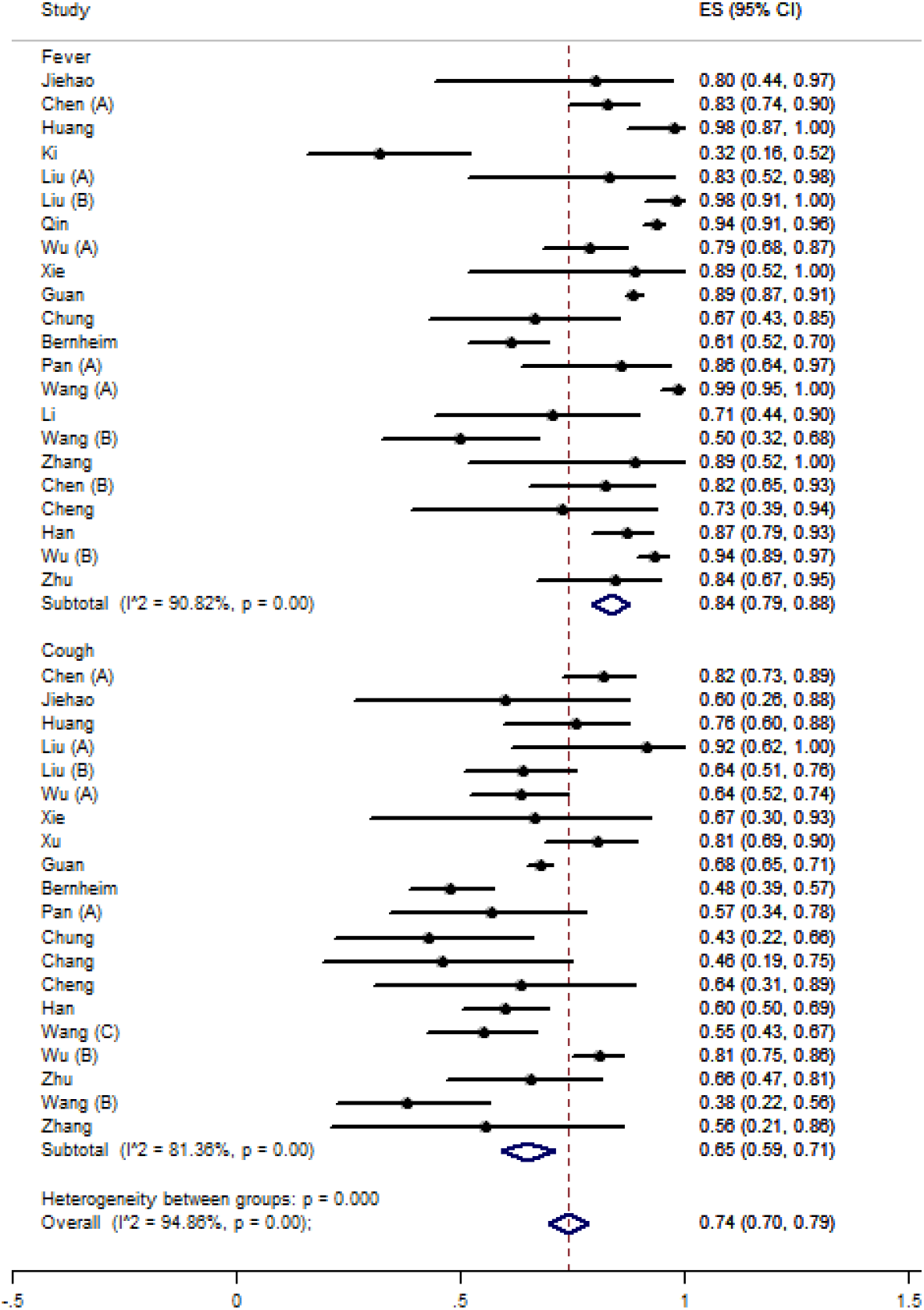
Meta-analysis of incidence of clinical findings of patients (A)

**Figure 5.**
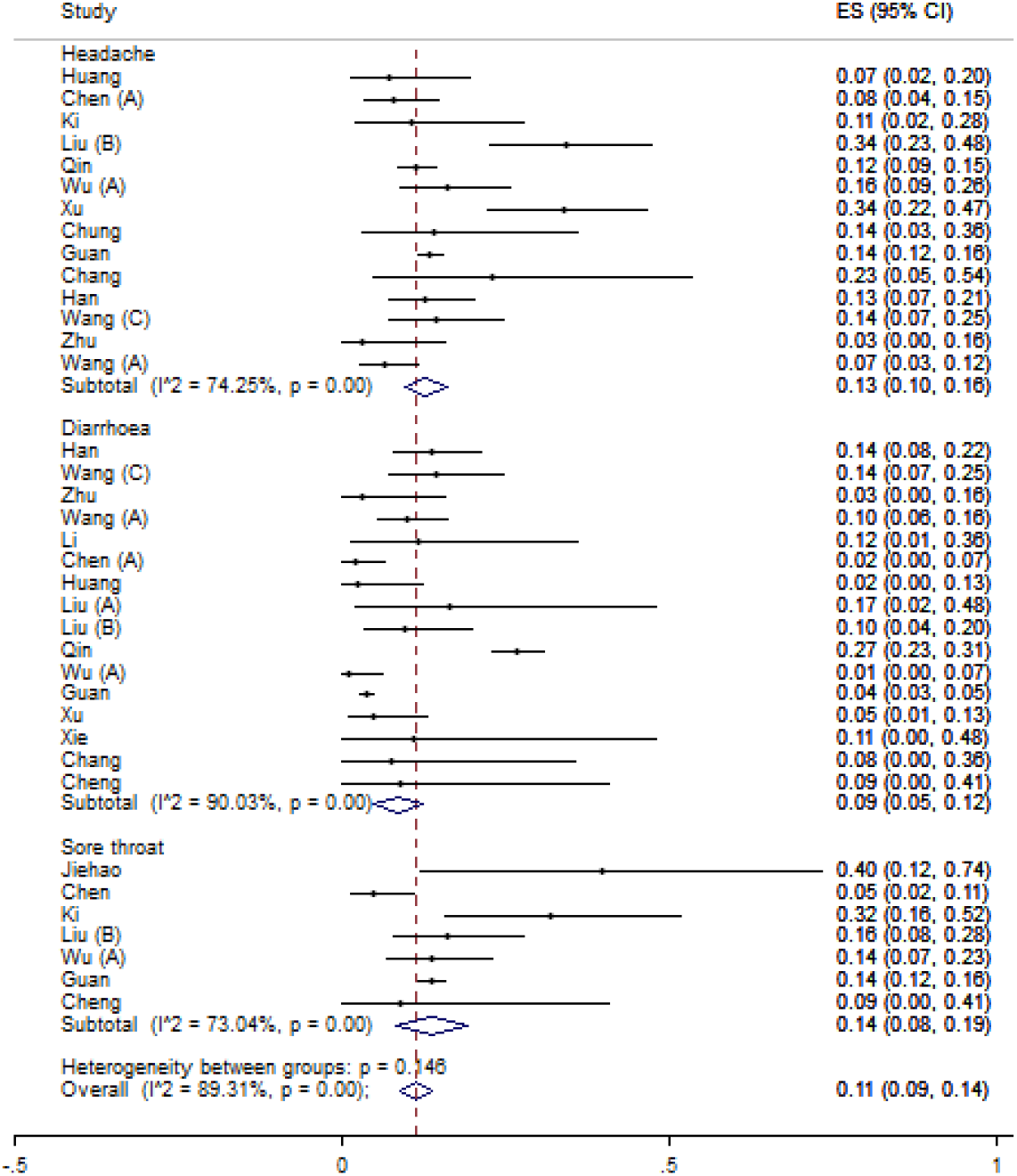
Meta-analysis of incidence of clinical findings of patients (B)

Headache, myalgia, diarrhea, and fatigue were reported in 14, 17, 16, and 13 studies with the risk of 13% (95% CI, 8-19), 24% (95% CI 19-29), 9% (95% CI, 5-12), and 42% (95% CI, 34-49) (Fig. 5 and Supplementary File). Anorexia, Chills, Shortness of breath, dyspnea, chest pain, confusion, and nausea/vomiting were reported in respectively 5, 5, 6, 5, 7, 2, and 9 studies. Overall prevalence of these symptoms was 24% (95% CI, 13-34), 19% (95% CI, 11-27), 27% (95% CI, 12-42), 31% (95% CI, 14-48), 6% (95% CI, 2-9), 1% (95% CI, 0-2) and 6% (95% CI, 3-8) (Supplementary File and Table 2).

**Table 2.** Summarized Pooled values of Considered findings

| Variable | All Patients |  |  | Severe |  |  | Non-severe |  |  |
| --- | --- | --- | --- | --- | --- | --- | --- | --- | --- |
|  | N. of study | I-squared | ES% (95%CI) | N. of study | I-squared | ES% (95%CI) | N. of study | I-squared | ES% (95%CI) |
| **Clinical outcomes** | - | - | - | - | - | - | - | - | - |
| Discharge | 5 | 96.58 | 26 (11 – 41) | - | - | - | - | - | - |
| Death | 6 | 77.22 | 3 (2 – 4) | - | - | - | - | - | - |
| Hospitalization | 5 | 98.24 | 69 (49 – 88) | - | - | - | - | - | - |
| **Clinical findings** | - | - | - | - | - | - | - | - | - |
| Fever | 22 | 90.82 | 84 (79 – 88) | 6 | 53.10 | 94 (90 – 97) | 6 | 92.97 | 90 (84 – 96) |
| Cough | 20 | 81.36 | 65 (59 – 71) | 6 | 48.60 | 75 (69 – 82) | 5 | 78.66 | 68 (60 – 76) |
| Sore throat | 7 | 73.04 | 14 (8 – 19) | 2 | - | 14 (9 – 19) | 2 | - | 14 (12 – 16) |
| Headache | 14 | 74.25 | 13 (10 – 16) | 5 | 39.32 | 12 (8 – 16) | 5 | 86.22 | 14 (8 – 20) |
| Myalgia | 17 | 82.18 | 24 (19 – 29) | 6 | 67.23 | 21 (14 – 28) | 5 | 88.09 | 27 (17 – 36) |
| Diarrhea | 16 | 90.03 | 9 (5 – 12) | 4 | 14.77 | 8 (3 – 12) | 5 | 73.49 | 9 (4 – 14) |
| Fatigue | 13 | 88.72 | 42 (34 – 49) | 6 | 88.70 | 53 (40 – 66) | 6 | 86.66 | 44 (35 – 53) |
| Anorexia | 5 | 89.13 | 24 (13 – 34) | 4 | 91.54 | 30 (9 – 51) | 4 | 78.88 | 17 (9 – 26) |
| Chills | 5 | 61.50 | 19 (11 – 27) | 2 | - | 16 (11 – 21) | 2 | - | 11 (9 – 13) |
| Shortness of breath | 6 | 97..16 | 27 (12 – 42) | 3 | - | 39 (20 – 59) | 3 | - | 19 (7 – 32) |
| Dyspnea | 5 | 95.41 | 31 (14 – 48) | 6 | 87.78 | 59 (41 – 76) | 4 | 2.45 | 24 (19 – 29) |
| Chest pain | 7 | 68.44 | 6 (2 – 9) | 2 | - | 2 (-1 – 6) | 2 | - | 4 (0 - 7) |
| Confusion | 2 | - | 1 (0 – 2) | - | - | - | - | - | - |
| Nausea/Vomiting | 9 | 81.88 | 6 (3 – 8) | 6 | 55.44 | 9 (5 – 13) | 4 | 42.88 | 6 (4 – 9) |
| Sneezing | 3 | - | 26 (14 – 39) | - | - | - | - | - | - |
| Nasal congestion | 4 | 85.68 | 24 (1 – 48) | - | - | - | - | - | - |
| Rhinorrhea | 7 | 27.70 | 4 (1 – 6) | 2 | - | 2 (1 – 4) | - | - | - |
| Hemoptysis | 3 | - | 2 (0 – 3) | 3 | - | 3 (1 – 5) | 3 | 0 | 1 (0 – 1) |
| Expectoration | 4 | 85.52 | 39 (27 – 51) | 2 | - | 39 (34 – 45) | 2 | 0 | 36 (30 – 41) |
| Abdominal pain | 2 | - | 4 (2 – 5) | 2 | 0 | 7 (4 – 10) | 2 | - | 6 (3 – 8) |
| Pharyngalgia | 5 | 83.00 | 12 (5 – 19) | 3 | - | 14 (-2 – 29) | 3 | - | 8 (5 – 12) |
| Dizziness | 4 | 00.00 | 8 (6 – 10) | 3 | - | 12 (5 – 18) | 3 | - | 5 (3 – 8) |
| Sputum production | 8 | 78.20 | 27 (20 – 35) | 4 | 53.39 | 41 (27 – 54) | 4 | 0 | 33 (30 – 36) |
| **Laboratory findings** | - | - | - | - | - | - | - | - | - |
| Leukocytosis | 10 | 82.05 | 8 (4 – 12) |  |  |  |  |  |  |
| Leukocytopenia | 13 | 95.29 | 26 (17 – 35) | - | - | - | - | - | - |
| Lymphocytopenia | 5 | 96.87 | 34 (12 – 57) | - | - | - | - | - | - |
| Neutrophilia | 4 | 82.16 | 25 (11 – 39) | - | - | - | - | - | - |
| Thrombocytopenia | 3 | - | 36 (30 – 42) | - | - | - | - | - | - |
| Thrombocytosis | 2 | - | 4 (1 – 8) | - | - | - | - | - | - |
| Increase in C-Reactive Protein | 8 | 99.34 | 56 (21 – 92) | - | - | - | - | - | - |
| Increase in Procalcitonin | 6 | 58.22 | 5 (1 – 8) | - | - | - | - | - | - |
| Increase in D-dimer | 4 | 92.14 | 14 (0 – 28) | - | - | - | - | - | - |
| Increase in ESR | 5 | 96.60 | 54 (28 – 79) | - | - | - | - | - | - |
| Increase in IL-6 | 2 | - | 42 (34 – 50) |  |  |  |  |  |  |
| **Radiologic findings** | 5 | 74.69 | 84 (79 – 88) | - | - | - | - | - | - |
| Ground-glass opacity | 11 | 99.52 | 50 (29 – 70) | - | - | - | - | - | - |
| Consolidation | 7 | 97.29 | 27 (10 – 44) | - | - | - | - | - | - |
| Unilateral pneumonia | 5 | 98.58 | 24 (3 – 44) | - | - | - | - | - | - |
| Bilateral pneumonia | 9 | 99.77 | 61 (30 – 91) | - | - | - | - | - | - |
| **Comorbidities** | - | - | - | - | - | - | - | - | - |
| Cardiovascular disease | 12 | 92.35 | 10 (7 – 114) | 5 | 70.43 | 15 (5 – 26) | 5 | 66.11 | 4 (1 – 6) |
| Cerebrovascular disease | 4 | 44.55 | 2 (1 – 3) | 3 | - | 10 (0 – 20) | 3 | - | 1 (1 – 2) |
| Diabetes | 11 | 79.17 | 10 (7 – 14) | 7 | 4.03 | 17 (14 – 21) | 7 | 67.85 | 6 (4 – 9) |
| Hypertension | 11 | 84.54 | 18 (13 – 23) | 5 | 76.93 | 31 (19 – 44) | 6 | 46.20 | 14 (11 – 18) |
| Chronic kidney disease | 7 | 30.24 | 1 (1 – 2) | 3 | - | 24 (-3 – 51) | 3 | - | 1 (0 – 2) |
| Digestive system disease | 3 | - | 5 (1 – 10) | - | - | - | - | - | - |
| Chronic liver disease | 8 | 33.07 | 2 (1 – 4) | - | - | - | 3 | - | 2 (1 – 4) |
| Endocrine system disease | 3 | - | 6 (-1 – 13) | - | - | - | - | - | - |
| Malignancy | 9 | 60.45 | 2 (1 – 3) | 3 | - | 2 (0 – 4) | 3 | - | 2 (0 – 3) |
| CNS disorder | 4 | 5.30 | 2 (1 – 4) | - | - | - | - | - | - |
| Respiratory system disease | 13 | 41.45 | 2 (1 – 3) | 5 | 21.15 | 6 (2 – 10) | 5 | 0 | 1 (0 – 1) |
| Immunodeficiency | 2 | - | 0 | - | - | - | - | - | - |
| **Complications** | - | - | - | - | - | - | - | - | - |
| Acute kidney injury | 3 | - | 2 (-1 – 5) | - | - | - | - | - | - |
| Acute respiratory distress syndrome | 4 | 91.70 | 21 (6 – 35) | - | - | - | - | - | - |

In addition, sneezing, nasal congestion, rhinorrhea, hemoptysis, and expectoration were reported in 3, 4, 7, 3, and 4 studies, respectively. Pooling results were 26% (95% CI, 14-39), 24% (95% CI, 1-48), 4% (95% CI 1-6), 2% (95% CI, 0-3), and 39% (95% CI, 27-51) (Supplementary File and Table 2).

Abdominal pain, pharyngula, dizziness, and sputum production have been reported in 2, 5, 4, 8 studies, respectively. Meta-analysis findings were 4% (95%CI 2 – 5), 12% (95%CI 5 – 19), 8% (95%CI 6 – 10), and 27% (95%CI 20 – 35) (Supplementary File and Table 2).

### Laboratory Findings

Many studies have investigated the laboratory findings of the patients. Most common observing in these patients were leukocytopenia (26%, 95%CI 17 – 35), neutrophilia (25%, 95%CI 11 – 39), thrombocytopenia (36%, 95%CI 30-42) and lymphocytopenia (34%, 95%CI 12 – 57) by following thrombocytosis (4%, 95%CI 1 – 8), increased C-reactive protein (CRP) (56%, 95%CI 21 – 92), procalcitonin (5%, 95%CI 1 – 8), IL-6 (42%, 95%CI 34 – 50), D-dimer (14%, 95%CI 0 – 28), and Erythrocyte sedimentation rate (ESR) (54%, 95%CI 28 – 79) (Fig. 7 and 8).

**Figure 6.**
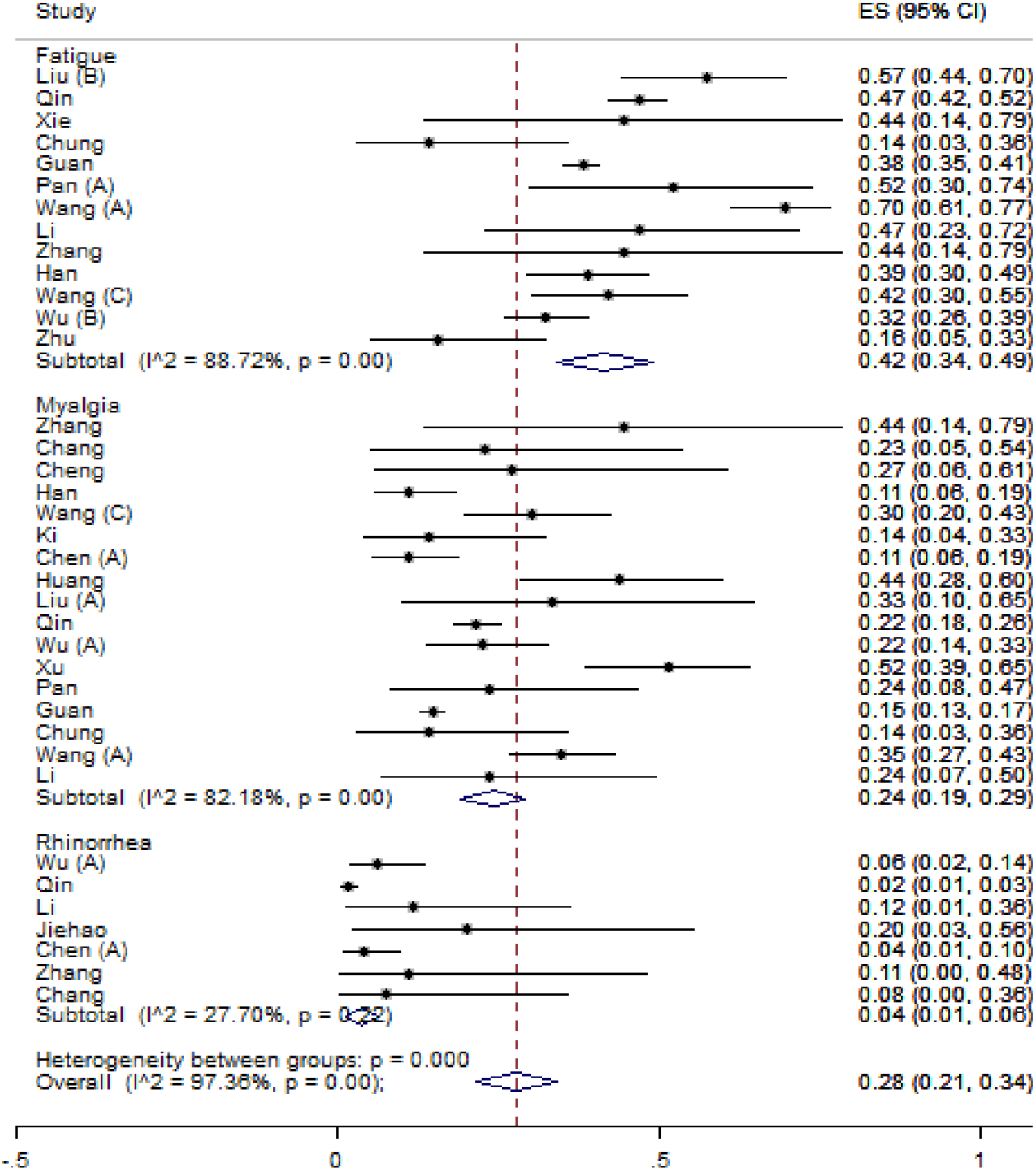
Meta-analysis of incidence of clinical findings of patients (C)

**Figure 7.**
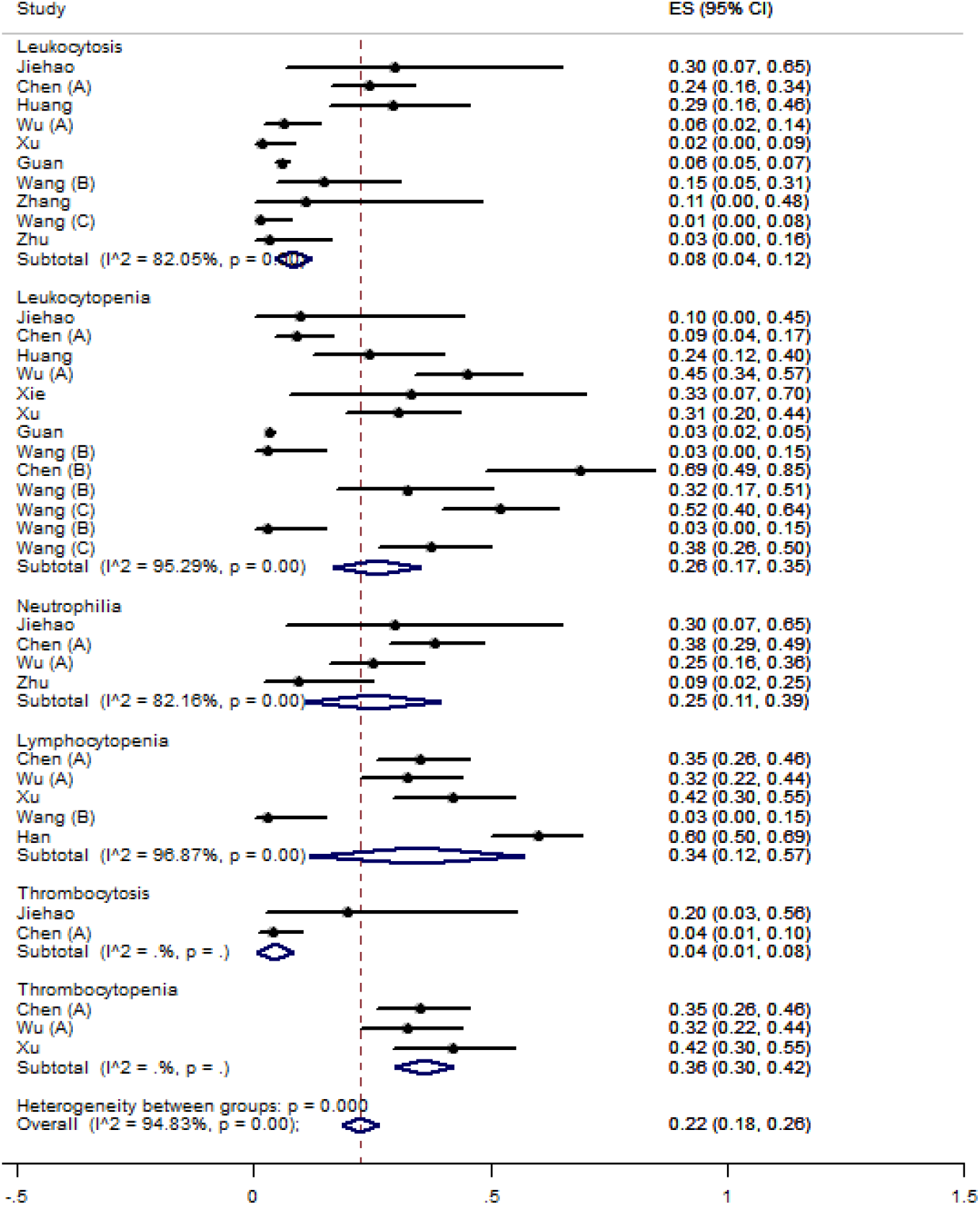
Meta-analysis of incidence of laboratory findings of patients (A)

**Figure 8.**
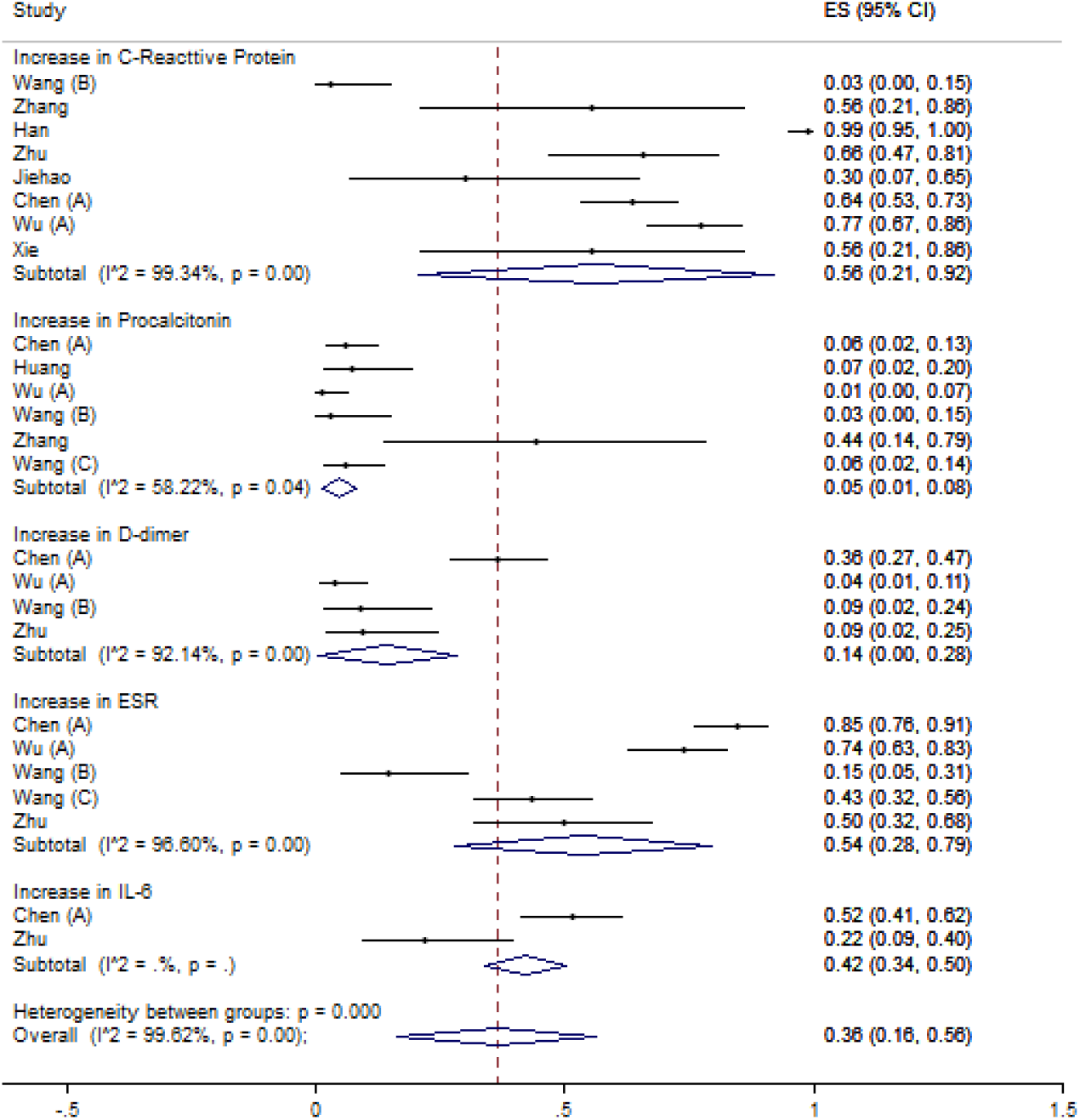
Meta-analysis of incidence of laboratory findings of patients (B)

### Radiological Findings

Regarding radiological findings, 84% (95%CI 79 – 88) of patients who underwent a Computed tomography (CT) scan were positive for pneumonia. The most common radiological findings in these patients were Ground-glass opacity (50%, 95%CI 29 – 70), consolidation (27%, 95%CI 10 – 44), unilateral pneumonia (24%, 95%CI 3 – 44), and bilateral pneumonia (61%, 95%CI 30 – 91) (Supplementary File and Table 2).

### Comorbidities

Different studies investigated the underlying diseases in patients with COVID-19. The cardiovascular disease, diabetes, and cerebrovascular disease were reported in 12, 11, and 4 original studies, respectively. In this regard, pooling of the results was 10% (95%CI 7 – 114), 10% (95%CI 7 – 14), and 2% (95%CI 1 – 3), respectively. Also, other underlying diseases like hypertension and chronic kidney disease were investigated in 11 and 7 studies. The general outbreak of these disorders was 18% (95%CI 13 – 23) and 1% (95%CI 1 – 2) respectively (Supplementary File and Table 2).

Other disorders prevalence was as follows: digestive tract (5%, 95%CI 1 – 10), chronic liver disease (2%, 95%CI 1 – 4), endocrine disease (6%, 95%CI -1 – 13), malignancies (2%, 95%CI 1 – 3), central nervous system (2%, 95%CI 1 – 4), respiratory system (2%, 95% 1 – 3) and immunodeficiency (0%) (Supplementary File and Table 2).

The acute respiratory distress syndrome and acute kidney injury after infecting with COVID-19 have been investigated in 4 and 3 studies, respectively. Meta-analysis showed the prevalence of 21% (95%CI 6 – 35) and 2% (95%CI -1-5) respectively (Supplementary File and Table 2).

### Comorbidities and clinical findings based on severity of disease

In this systematic review for detecting the source of heterogeneity, we analyzed the subgroups based on the severity of the disease. The results are as follows:

The prevalence of the symptoms such as sore throat, sputum production, headache, and fatigue in stable patients did not need to be admitted into intensive care unit (ICU) were 14%, 33%, 14%, 44%, respectively. However, these statistics for patients with severe symptoms were 14%, 41%, 12%, 53%. Other symptoms like diarrhea, anorexia, nausea/vomiting, and dyspnea in mild patients showed the prevalence of 9%, 17%, 6%, and 24%, which were 8%, 30%, 9%, and 59%, respectively in severe cases (Supplementary File and Table 2).

The prevalence of symptoms like abdominal pain, pharyngalgia, hemoptysis, expectoration, and fever in mild cases were 6%, 8%, 1%, 36%, 90%. The same symptoms for the severe cases were 7%, 14%, 3%, 39%, 94% respectively. Furthermore, the prevalence of the symptoms of myalgia, cough, dizziness, chills, and shortness of breath were 27%, 68%, 5%, 11%, and 19% whereas in severe cases were 21%, 75%, 12%, 16%, and 39%, respectively (Supplementary File and Table 2).

The meta-analysis showed that the prevalence of the underlying diseases like cardiovascular disease, diabetes, cerebrovascular disease and hypertension in COVID-19 patients were 4%, 6%, 1%, 14%, respectively. However, in critically severe patients were 15%, 17%, 10%, and 31%. The prevalence of other underlying diseases like chronic liver disease and respiratory diseases were 2% and 1% whereas in severe cases, respiratory diseases prevalence is 6% (Supplementary File and Table 2).

The prevalence of malignancies (2%) and chronic kidney disease (1%) mostly reported in mild patients whereas in severe cases, were 2% and 24%, respectively (Supplementary File and Table 2).

## Discussion

Due to the novelty of Coronavirus 2019, there is no clear picture of the clinical and paraclinical features of the disease. Moreover, the reported frequencies of these features are variable. Therefore, this systematic review and meta-analysis study was performed to evaluate the clinical and paraclinical features of the disease. In this systematic review, 20 studies including 48967 cases of COVID-19 have been studied. All the studies were conducted in China, with one exception which studies some case in South Korea along with Chinese cases.

It was difficult to diagnose this disease at the time of onset, as the general symptoms of the disease can also be seen in other respiratory diseases. A wide range of clinical, laboratory, and imaging findings have been observed in relation to this disease. Symptoms such as fever, cough, fatigue, and sputum were common clinical symptoms in this study, but cases such as diarrhea, chest pain and nausea were less common. As mentioned, gastrointestinal symptoms such as diarrhea and nausea are less common and symptoms such as fever and cough are more common in viral infections such as seasonal influenza, SARS and MERS. (8, 9). Many patients with coronavirus may be unrecognized and present in the community because, according to the results of this study, none of the clinical symptoms were definitively present in all patients. Therefore, definitive diagnosis of patients is difficult and one should expect hidden and vector-borne patients in the community. In the meta-analysis of Sun *et al*. which conducted on the symptoms of COVID-19 patients, the results have shown that fever, cough, and fatigue are among the most common symptoms (7).

The rate of death and the outcome of patients after hospitalization have been investigated in this meta-analysis. The results have shown more than two-thirds of the patients (69%) who referred to the hospitals get hospitalized, and about 26% will be released after outpatient treatment. Around 3% of the patients, however, will expire. It should be noted that the real mortality rate would be higher than this statistics. In the study of Baud *et al*. It has been shown that the mortality of this disease is higher than that typically obtained by dividing the death rate by the total number of patients. Because at the time of infection the number of persons is much lower than the number entered into the denominator (10). Based on the present meta-analysis findings in 84% of suspected cases, CT-Scan findings were positive, indicating that a high proportion of patients can be identified by relying on CT-Scan.

On the other hand, 84% of suspected cases had high fever and cough was seen in many patients. Changes in laboratory findings are less common than radiological and clinical findings and appear to be less reliable. According to the study in the group of patients with severe conditions and outpatients, it was found that the symptoms associated with the disease are more common in critically ill patients at the time of diagnosis and treatment. Also, underlying diseases were more common in these patients.

The limitations of this study include:

1. All of the studies have been conducted in China
2. High methodological heterogeneity in the included studies
3. Due to the lack of information by sex and age, it was not possible to calculate the mortality and morbidity of patients in these subgroups.

### Conclusion

Given the high proportion that may occur without CT-Scan findings or clinical symptoms, it is advisable to use several combination methods to better diagnose the disease, to minimize undiagnosed patients. Moreover, while there is no access to clinical and paraclinical facilities in in public places such as airports and border crossings, it is recommended to consider factors such as fever, cough, sputum and fatigue. Since the prevalence of underlying diseases is higher in patients with more severe conditions, the risk of serious illness in those with underlying diseases should be considered.

## Data Availability

The data that support the findings of this study are openly available in data bases mentioned in the search strategy.

## Acknowledgment

The authors would like to thank the Student Research Committee of Mazandaran University of Medical Sciences for supporting this project (Project No. 7319).

## Conflict of interest

The authors have no conflicts of interest to declare.

## Supporting Information Legend

Supplementary figure 1. Risk of bias summary (B)

Supplementary figure 2. Mete-analysis of incidence of Clinical findings of patients (A)

Supplementary figure 3. Mete-analysis of incidence of Clinical findings of patients (B)

Supplementary figure 4. Mete-analysis of incidence of Radiologic findings

Supplementary figure 5. Mete-analysis of prevalence of Comorbidities (A)

Supplementary figure 6. Mete-analysis of prevalence of Comorbidities (B)

Supplementary figure 7. Mete-analysis of prevalence of Clinical findings in severe patients (A)

Supplementary figure 8. Mete-analysis of prevalence of Clinical findings in severe patients (B)

Supplementary figure 9. Mete-analysis of prevalence of Clinical findings in non-severe patients (A)

Supplementary figure 10. Mete-analysis of prevalence of Clinical findings in non-severe patients (B)

Supplementary figure 11. Mete-analysis of prevalence of Comorbidities in severe patients

Supplementary figure 12. Mete-analysis of prevalence of Comorbidities in non-severe patients

## Notes

### Competing Interest Statement

The authors have declared no competing interest.

### Funding Statement

We have no funds for this study.

